# Anxiety appraisal in mothers of preterm neonates admitted in critical care unit

**DOI:** 10.1101/2024.01.10.24301141

**Authors:** Rafia Gul, Samer Fatima, Samina Khurshid, Sidra Niamat, Zahid Anwar, Saher Gul Ahdi

## Abstract

**Background:** Mothers of premature neonates often confront various psychological challenges including postpartum depression, anxiety, and elevated stress levels. However, anxiety has not received the necessary emphasis in routine clinical practice and research, often going unnoticed. There is insufficient data regarding the utilization of specific tools for screening maternal anxiety in hospitalized preterm infants. The study aimed to determine the prevalence of maternal anxiety and its risk factors among mothers of premature neonates admitted to the intensive care unit.

**Methods:** Following Institutional Review Board (IRB) approval, a descriptive cross-sectional study was conducted at level-III Neonatal Unit, Fatima Memorial Hospital Shadman, Lahore over duration of 30 months, (January 2021-July 2023). Mothers werescreened for anxiety using Perinatal Anxiety Screening Scale (PASS) having 31 items. Comparative descriptive statistics and multiple logistic regression were applied to identify all risk factor while taking p <0.05 as significant.

**Results:** Out of 430 mothers, 28.1% experienced anxiety. Statistically significant factors for maternal anxiety include young age, urban residence, higher education, smoking, diabetes, depression/anxiety diagnosis, primiparity, delivery complications, maternal involvement in neonatal care, breastfeeding, fear of handling preterm infants at home, and prematurity-related concerns, as well as gestational age, birth weight, and weight for gestational age (p < 0.05). Among all these, however, prematurity <28 weeks (p < 0.001, AOR 496, 95% CI 22.5–10951) and 28-31+6 weeks (p < 0.001, AOR 265, 95% CI 14– 5010), alongwith primiparity (p = 0.001, AOR 16.483, 95% CI 3.287–82.648), and SGA (p = 0.025, AOR 8.9, 95% CI 1.3–60.6) increase while extended family system protects from maternal anxiety (p = 0.002, AOR 0.25, 95% CI 0.106–0.595).

**Conclusion:** Every third mother in our study population who delivered prematurely experienced anxiety. The younger gestational age, first-time motherhood, SGA neonate increase while the extended family system serves as a protective factor against maternal anxiety.

## Introduction

Preterm birth is the occurrence of a live birth before 37 weeks of gestation ^(1)^. Approximately every tenth newborn worldwide is born prematurely ^(2)^. Pakistan has a high incidence of preterm births, accounting for 15.8% of all births ^(3)^. Prematurity is traumatic to mothers as it has physiological and psychological effects in the form of emotional distress, depression, and/or anxiety among postpartum mothers ^(4)^.

Adding a child to the family is typically a rewarding experience for mothers, but it is always accompanied by a degree of anxiety. Postpartum excessive and debilitating maternal anxiety is a significant health issue. Its traumatic effects compromise maternal health and impede the development of mother-baby connections at a time when they are most sensitive to their environments, resulting in poorer behavioral, cognitive, and emotional development outcomes for neonates ^(5,6)^.

In the postpartum period, 3–43% of mothers have reported anxiety symptoms ^(7, 8)^. Maternal (demographic, socioeconomic, obstetric, previous experiences of having a premature baby, family support, severity of prematurity, and current diagnosis of depression or anxiety); neonatal (clinical status of the neonate); and mother-baby interaction are contributory factors for maternal anxiety during the postpartum period. (5) Globally, different instruments have been developed to assess and measure anxiety. However, only a few tools are available that can be used to o screen for maternal anxiety during the postpartum period. The Perinatal Anxiety Screening Scale (PASS) is an example of such an instrument that has been designed specifically for the perinatal period, this tool is user-friendly and condition-specific ^(9 – 11)^. Its evaluation imposes a negligible additional burden on mothers and health care. The PASS is easily applicable as it is shorter (31 items) than other instruments such as the STAI (State-Trait Anxiety Inventory) with 41 items.

Nevertheless, anxiety has received less attention than it deserves in routine clinical practice and research, remaining largely undetected and untreated. Accordingly, the body of literature, anxiety and depression are frequently reported interchangeably, and the majority of available research focuses on both maternal stress and depressive symptoms. Only the tip of the iceberg is known about maternal anxiety in the postpartum period. In addition, there is limited data regarding maternal screening for anxiety in hospitalized preterm infants using specific tools. According to the author’s best knowledge, no such local data exists ^(12 – 16)^. This study will aid in the evaluation of postpartum anxiety in mothers with a hospitalized premature infant.

The study’s objectives included addressing the frequency and severity of maternal anxiety of mothers of premature neonates admitted to the intensive care unit. In addition, the current study was conducted to determine the relationship between demographic, socioeconomic, obstetric, and other psychosocial variables and maternal anxiety.

## Material and methods

Following Institutional Review Board approval, a descriptive cross-sectional study was conducted at level-III Neonatal Unit, Fatima Memorial Hospital Shadman, Lahore over duration of 30 months, (January 2021 – July 2023). The inclusion criteria were neonates who were singletons, either inborn or referred from other hospitals for preterm care, and consent to participate in the study. All neonates with malformations, anomalies incompatible with life, death within three days of postnatal life, severely ill mothers, maternal unavailability, multifetal pregnancies, refusal to participate, or missing data were excluded.

## Data collection

We enrolled 504 maternal-neonatal pairs after obtaining the mothers’ written consent. The data were collected using the study tools, including a questionnaire on socio-demographic data, the Prenatal Anxiety Screening Scale (PASS), and researcher-made questions on maternal anxiety during the early postnatal period. The PASS questionnaire was designed by Somerville et al. for the screening of maternal anxiety and its validity and reliability were confirmed. The average time taken for respondents to complete the PASS is 6 minutes ^(12)^. We have taken 20 as the cut-off point as mothers with a score of >20 were labeled to have anxiety.

The mothers were interviewed in the early postpartum period, between three and twenty-one days after childbirth for anxiety appraisal using the PASS questionnaire. It is a straightforward four-factor configuration, as illustrated below:

a. Acute anxiety and adaptability (questions 1–10)
b. General anxiety and specific fears (questions 11–18)
c. Perfectionism, control, and trauma (questions 19–23)
d. Social anxiety (questions 24–31)

Scoring for each question is done using a 4-point Likert scale and it ranges from 0 to 3. Choosing the option not at all has a score of “0”, sometimes “1”, often almost “2” and always as”3”.

To score the questionnaire, the scores of the individual questions are added up, and a score of ≥26 is the cut-off point between low and high risk of anxiety. The recommended severity ranges depending on the score are as:

- 0–20: asymptomatic
- 21–41: mild anxiety
- 42–93: severe anxiety

The researcher-developed questionnaire was created by first conducting a comprehensive literature review and analyzing systematic reviews in the following areas: (i) psychological and psychiatric factors, (ii) factors concerning social support and marital relationships, (iii) social and demographic factors, (iv) factors related to pregnancy and its related problems, and (v) neonatal factors. Subsequently, an experienced panel of experts completed the questionnaires, ensuring that all items were extracted from the literature review and validated by their expertise. Following this, the questionnaire underwent a pilot study, with fifteen mothers participating to ensure the validity of the items.

The neonatal characteristics, including gender, weight, gestational age, birth weight, and weight for gestation were collected. Maternal characteristics include age, place of residence (city or village), level of education (uneducated, primary, secondary, or higher), employment status (housewife or employed), smoking, any drug addiction, hypertensive disorders of pregnancy, gestational diabetes mellitus, current diagnosis of depression or anxiety, parity, previous experience with preterm babies, mode of delivery (SVD or LSCS), family system (extended or nuclear), involved in baby care and breastfeeding, fear of complications related to prematurity, fear of difficulty in handling preterm at home, and post-delivery complications (postpartum hemorrhage, fever, and infections). In addition, the status of the spouse was enquired about, including whether he resides alongside or abroad, and whether he is currently accompanying or not.

## Data Analysis

The data were analyzed with SPSS 21. The normality of data was analyzed with the Shapiro-Wilk test. Frequencies and percentages were used for qualitative while medians and IQR were used for non-normal quantitative variables respectively. All categorical variables were analyzed by proportional differences with either the Pearson chi-square test or Fisher’s exact tests. The z-test was applied to compare column proportions, and p-values were adjusted using the Bonferroni method. Mann-Whitney test was used to analyze quantitative variables in the two non-normal groups. Independent risk factors for maternal anxiety were identified by comparing demographic data and clinical characteristics of mothers with and without anxiety. In addition, logistic regression analysis was used to analyze and calculate the effects of quantitative and qualitative data on anxiety. Statistical significance was set at 5%.

## Results

A total of 504 mothers were enrolled in the study after fulfilling the inclusion criteria. However, 430 (85.3%) mother-neonatal pairs were finally enrolled in the study and has been summarized in Figure 1

**Figure 1.**
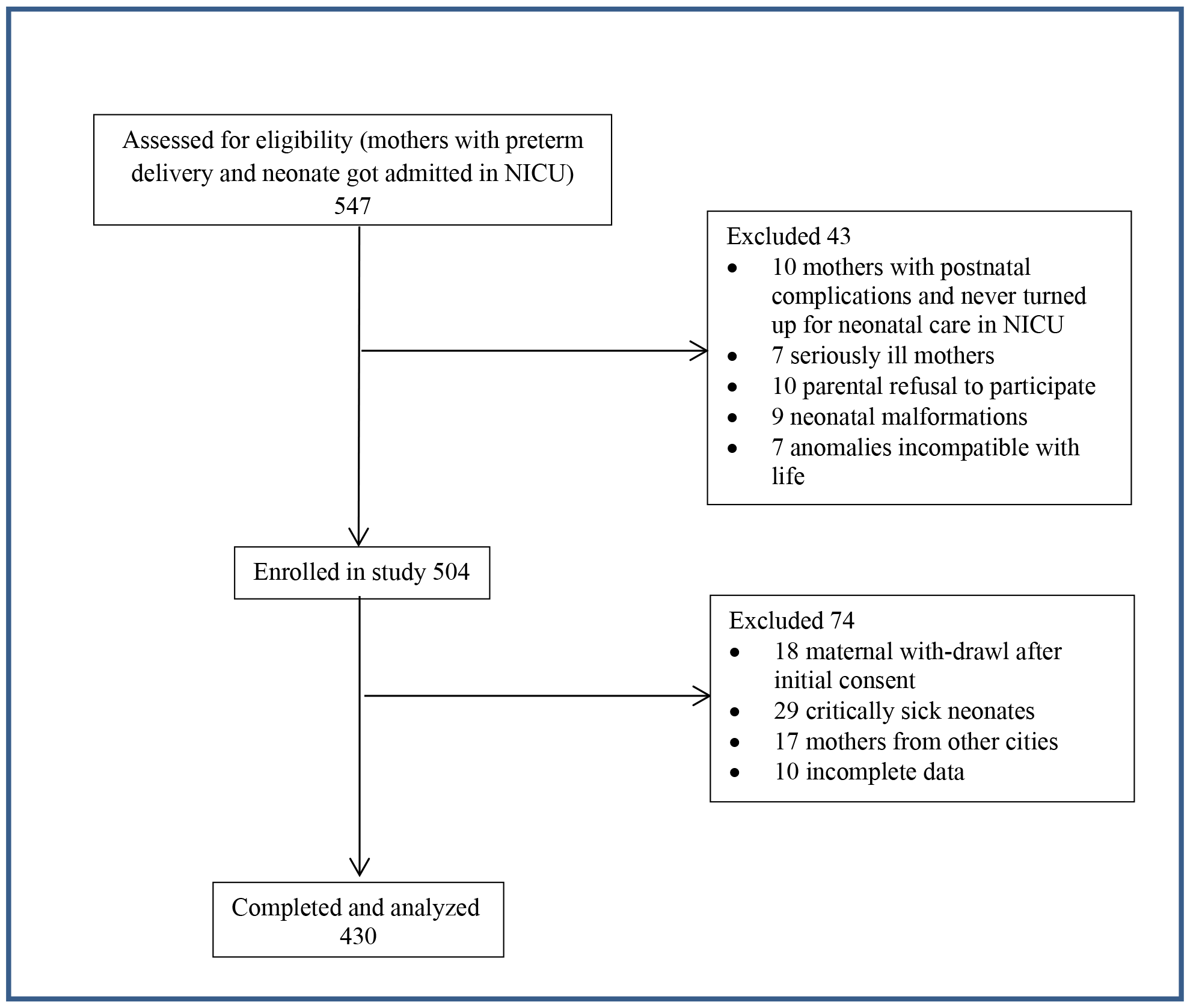
Consort diagram

All mothers were screened for their anxiety using the Perinatal Anxiety Screening Scale (PASS) scale and their response has been summarized in Figure 2.

**Figure 2.**
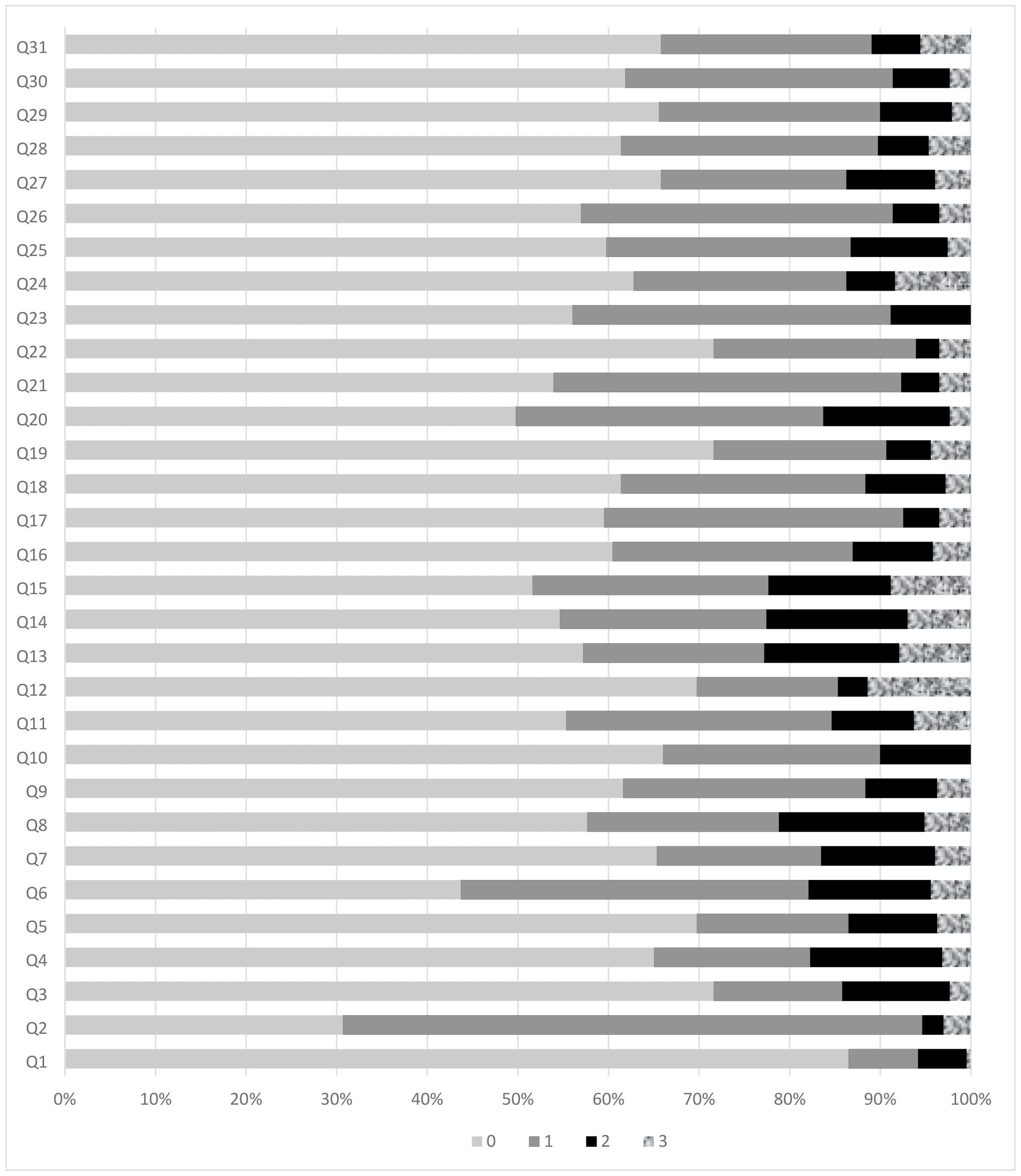
Maternal Response to the Perinatal Anxiety Screening Scale (“0”, sometimes “1”, often almost “2” and always as”3”)

Out of 430 mothers, 121 (28.1%) had anxiety with 19% mild to moderate and 9% with sever anxiety.

Table 1summarized demographic data of mothers and neonates. Of all the demographic and background variables of the study, young maternal age (p = 0.004), urban residence (p = 0.008), secondary and higher education level (p = <0.001), smoking (p = 0.049), diabetes mellitus (p = <0.001), current diagnosis of depression/anxiety (p = 0.007), primiparity (p = <0.001), mode of delivery (p = 0.038), post-delivery problems (p = 0.003), mothers involved in baby care (p= 0.047), breastfeeding / providing expressed breast milk (EBM) for neonate (p = 0.004), fear of difficulty in handling preterm at home (p = 0.018, maternal fear of prematurity related problems (p = 0.005), extended family system (p = 0.006), gestation age (p = <0.001), birth weight (p = <0.001), and weight for gestation age (p = 0.034). However, the variables of maternal job status, drug addiction, spouse living abroad, accompanied by a spouse, and gender of neonate were not associated with anxiety (p > 0.05) (Table 1).

**Table 1:**
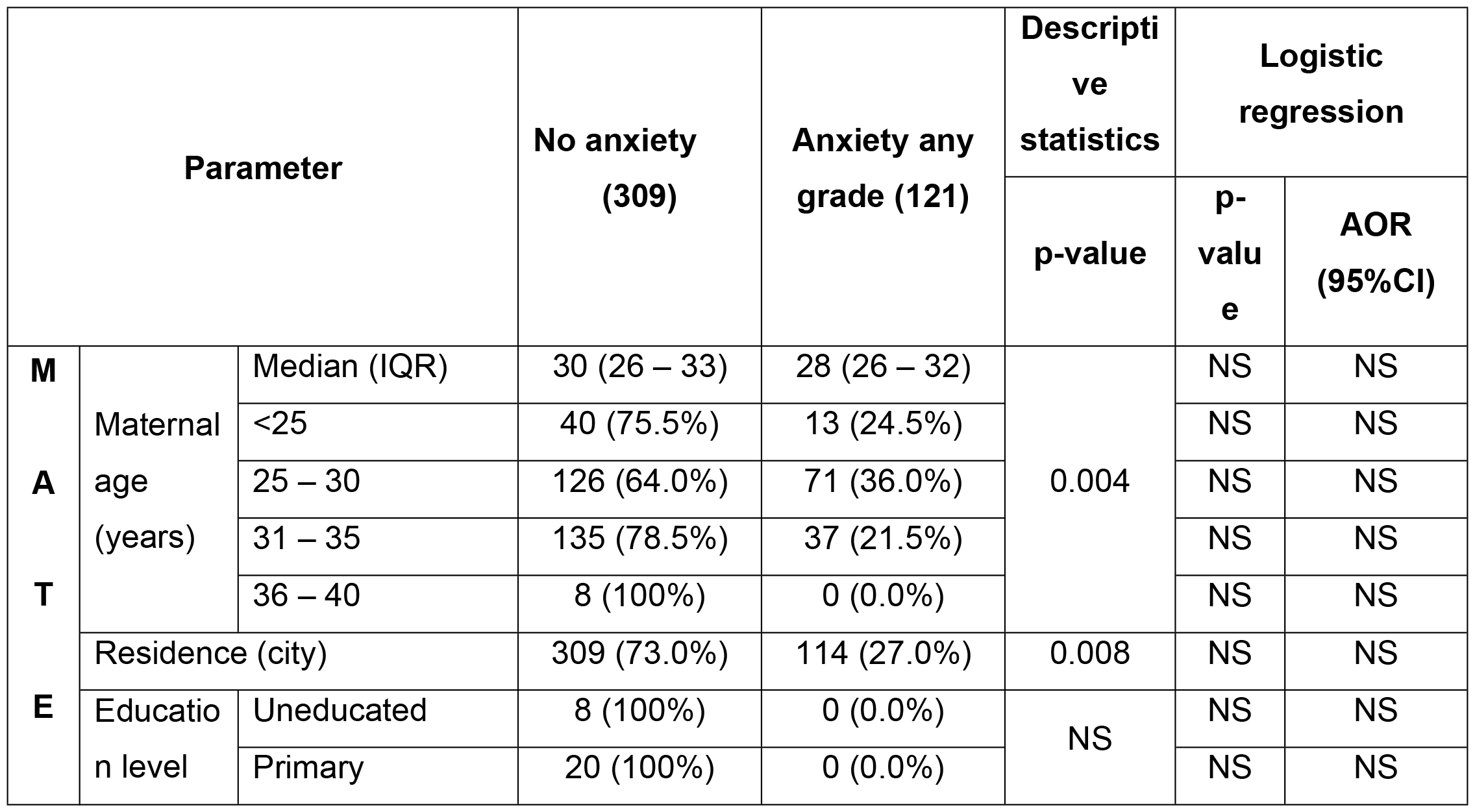
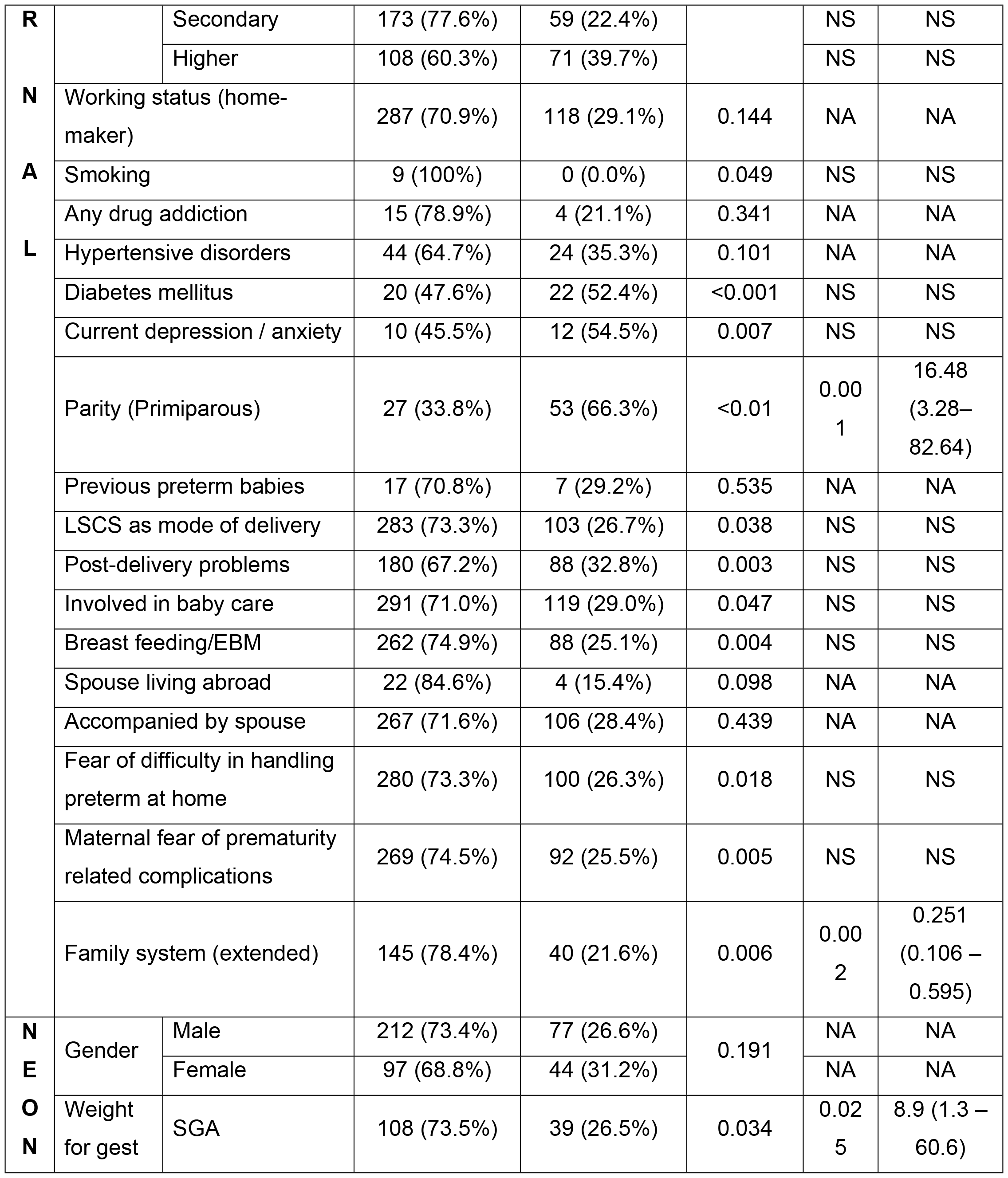
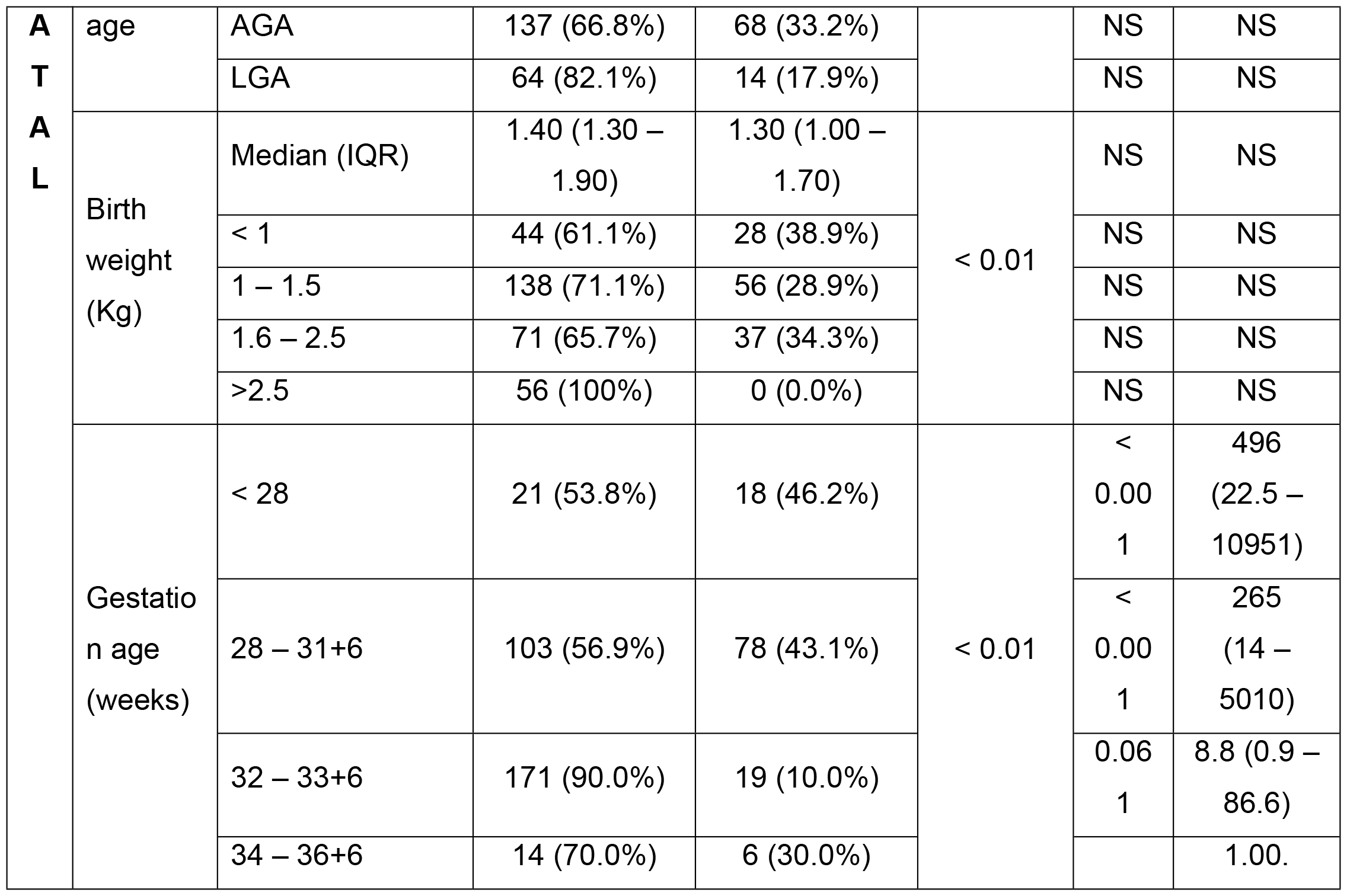
Risk factors of maternal anxiety.

The logistic regression analysis, as shown in Table 1, ultimately revealed that, after accounting for other variables, postnatal anxiety was linked to factors such as having a preterm neonate with a gestational age of less than 32 weeks, being small for gestational age (SGA), being a first-time mother (primiparous), and residing in an extended family system. The most significant risk of anxiety was observed in mothers whose babies were born preterm at a gestational age of less than 28 weeks, with a substantial increase of 496%. Next in line were mothers who gave birth within the gestational age range of 28 to 31+6 weeks, experiencing a heightened risk increase of 265%. Primiparous mothers were found to be 16% more likely to experience anxiety compared to multiparous mothers (p = 0.025). Furthermore, mothers whose infants were both preterm and small for gestational age (SGA) faced an increased risk of anxiety, with an 8.9% higher likelihood. On the flip side, the extended family system had a protective influence, decreasing anxiety by 75% in mothers when compared to those residing in nuclear family setups.

## Discussion

The journey to motherhood can be challenging due to the physical, emotional, and behavioral changes that occur during pregnancy and the postpartum period. This transition becomes even more intricate and demanding when a premature baby is born. Among the psychological challenges faced by mothers of premature neonates, anxiety stands out as a particularly prevalent issue. The present study aimed to evaluate the prevalence and risk factors related to anxiety of mothers of premature neonates admitted to the intensive care unit.

Our study signposts that approximately one in three mothers with preterm neonates in the NICU reported experiencing anxiety. These findings align closely with the prevalence reported by Benner and Cenna with reported prevalence as 26.5% and 34.2% respectively ^(13, 14)^. In contrast, Dannis et al. and Radosn et al. reported a significantly lower prevalence of anxiety, approximately 50% less than what we observed in our study ^(12, 15)^.

Variations in the prevalence of maternal postnatal anxiety, which persisted even when utilizing the same diagnostic tool, may be attributed to (i) specific individual and clinical characteristics of the mothers, (ii) the level of maternal engagement in neonatal care and breastfeeding, and (iii) the presence of drop-outs. It’s worth noting that a significant reason for individuals declining to participate in our study was that some women felt they were in good health and had confidence that they wouldn’t experience anxiety.

The PASS instrument offers valuable attributes, including its capacity for distinguishing and predicting accurately, its reliability over short-term test-retest intervals, user-friendliness, widespread acceptance, and the ability to facilitate precise cross-country comparisons. These aspects collectively enhance its ability to provide a reasonably reliable estimate of anxiety prevalence when applied ^(12, 16)^.

Our research findings revealed a substantial correlation between gestational age and maternal anxiety. As gestational age decreases, maternal anxiety tends to increase, and vice versa. In our study’s participant pool, giving birth at 28 weeks or less was associated with a 496-fold increase in maternal anxiety, while in the gestational age group of 28 to 32 weeks, the increase in maternal anxiety was 265-fold. Gonzalez-Hernandez and co found that about 33% of Mexican mothers who delivered prematurely experienced anxiety ^(16)^. However, Dantas et al. and Ong et al. reported that every 8^th^ woman with preterm neonate experiences anxiety ^(!2,17)^. The unusually high prevalence of anxiety observed in these studies can be attributed to (i) varying diagnostic criteria, (ii) the level of prematurity in newborns, and (iii) apprehension about complications associated with prematurity.

Additionally, we found that primiparity significantly increased the risk of postnatal maternal anxiety, supporting earlier research that linked anxiety symptoms to first-time mothers ^(14)^. It is generally accepted that multiparous mothers have more experience, training, and expertise in parenting, all of which are protective factors against anxiety ^(18)^. However, our findings contradict certain prior research results that suggested an association between anxiety symptoms and multiparity, but notably, such an association only appeared among multiparous women who also had a psychiatric history or experienced high levels of stress ^(19)^.

We have discovered that when prematurity occurs alongside small for gestational age (SGA) status, it amplifies the burden of maternal anxiety. This heightened anxiety may be attributed to maternal concerns about additional challenges linked to SGA, including a higher risk of infections, hypoglycemia, feeding difficulties, and various other potential issues. International data on the correlation between maternal anxiety and SGA neonates is limited.

The present study shows that the extended family system served as a protective factor against postnatal anxiety. There is literary no available data in the targeted population of mothers with preterm neonates admitted in NICU. However, in general, it has been reported that adolescents from joint families have better mental health compared to nuclear families ^(20, 21)^.

The extended family system can provide valuable support to mothers, offering emotional assistance, practical aid, help with childcare, knowledge exchange, adaptability in familial roles, and alleviation of feelings of isolation, ultimately leading to a reduction in maternal anxiety ^(5, 6)^.

In our study population, every second pregnant woman with gestational diabetes was diagnosed to have anxiety. Gestational diabetes can be distressing for mothers, affecting their mental well-being during and after pregnancy, especially in the setting of a neonate who is premature and admitted to the NICU. Other research findings corroborate our observation of a varying degree of association between gestational diabetes and maternal anxiety ^(19 – 22)^.

The present study showed that young maternal age, higher education, urban residence, current diagnosis of anxiety or depression, LSCS as the mode of delivery, post-partum complications, having a low birth weight baby, and maternal involvement in neonatal care and breastfeeding increased the risk of postnatal anxiety symptoms confirming previous findings ^(11 – 15, 22 – 25)^.

Maternal fear of complications related to prematurity and handling preterm neonates at home were the other significant risk factors for maternal anxiety but could not be compared as, to our knowledge, they were not included in comparable studies.

## Conclusion

What is known about postpartum anxiety in mothers is only the tip of the iceberg. Postpartum anxiety is unseen but common. In the postpartum period, maternal anxiety increases in response to real-world stressors, and it multiplies in the presence of preterm birth. Approximately 30% of mothers who delivered prematurely experienced anxiety, with the most significant risk factors being a younger gestational age, first-time motherhood, and having a baby who is small for their gestational age. Notably, the extended family system played a protective role against maternal anxiety.

## Data Availability

All relevant data are within the manuscript and its Supporting Information files.

## Acknowledgement

Mr. Abdul Basit Mubeen

## References

1. Preterm birth. Available at: https://www.who.int/news-room/fact-sheets/detail/pretermbirth.

2. Walani SR. Global burden of preterm birth. Int J Gynecol Obstet 2020;150:31–33

3. Every preemie-scale Pakistan 2016. Available at http://www.EveryPreemie.org

4. Mizrak B, Deniz AO, Acikgoz A. Anxiety levels of mothers with newborns in a neonatal intensive care unit in Turkey. Pak J Med Sci 2015;31:1176–81. doi: 10.12669/pjms.315.7792. PMID: 26649009; PMCID: PMC4641278.

5. Celen R, Arslan FT. The anxiety levels of the parents of premature infants and related factors. J Pediatr Res 2017;4(2):68–74. DOI: 10.4274/jpr.65882

6. Johnson K. Maternal-infant bonding: A review of literature. International Journal of Childbirth Education. 2013;28(3);17–22.

7. Segre LS, McCabe JE, Chuffo-Siewert R, O’Hara MW. Depression and anxiety symptoms in mothers of newborns hospitalized on the neonatal intensive care unit. Nursing Research 2014;63(5):320–332. PMID:25171558; PMCID: PMC4151274

8. McCabe-Beane JE, Stasik-O’Brien SM, Segre LS. Anxiety screening during assessment of emotional distress in mothers of hospitalized newborns. JOGNN 2018;47:105–113. 10.1016/j.jogn.2017.01.013. PMID: 28528808.

9. Fallon V, Halford JCG, Bennett KM, Harrold JA. The Postpartum Specific Anxiety Scale: development and preliminary validation. Arch Womens Ment Health 2016;19:1079–1090. DOI 10.1007/s00737-016-0658-9. PMID:27571782; PMCID: PMC5102940.

10. Bingol BF, Bal MD, Ozkan SA, Zengin O, Civ B. The adaptation of the Postpartum-Specific Anxiety Scale into the Turkish language. J Reprod Infant Psychol 2019;1–14. doi:10.1080/02646838.2019.1705265. PMID:31870187.

11. Fallon V, Silverio SA, Halford JCG, Bennett KM, Harrold JA. Postpartum-specific anxiety and maternal bonding: Further evidence to support the use of childbearing specific mood tools. J Reprod Infant Psychol 2021;39(2):114–124. doi: 10.1080/02646838.2019.1680960. Epub 2019 Oct 23. PMID:31642692

12. Dennis CL, Falah-Hassani K, Shiri R. Prevalence of antenatal and postnatal anxiety: systematic review and meta-analysis. Br J Psychiatry 2017;210(5):315–323. doi: 10.1192/bjp.bp.116.187179. Epub 2017 Mar 16. PMID:28302701

13. Bener A. Psychological distress among postpartum mothers of preterm infants and associated factors: a neglected public health problem. Revista Brasileira de Psiquiatria 2013;35:231–236. doi:10.1590/1516-4446-2012-082117

14. Cena L, Gigantesco A, Mirabella F, Palumbo G, Trainini A, Stefana A. Prevalence of maternal postnatal anxiety and its association with demographic and socioeconomic factors: A multicentre study in Italy. Front Psychiatry 2021;12:737666. doi: 10.3389/fpsyt.2021.737666. PMID:34658970; PMCID: PMC8514655

15. Rados NS, Tadinac M, Herman R. Anxiety during pregnancy and postpartum: Course, predictors and comorbidity with postpartum depression. Acta Clin Croat 2018;57(1):39–51. doi: 10.20471/acc.2017.56.04.05. PMID:30256010; PMCID: PMC6400346.

16. Gonzalez-Hernandez A, Gonzalez-Hernandez D, Fortuny-Falconi CM, Tovilla-Zárate CA, Fresan A, Nolasco-Rosales GA, et al. Prevalence and Associated Factors to Depression and Anxiety in Women with Premature Babies Hospitalized in a Neonatal Intensive-Care Unit in a Mexican Population. J Pediatr Nurs 2019;45:e53–e56. doi: 10.1016/j.pedn.2019.01.004. Epub 2019 Jan 14. PMID:30655115

17. Ong SL, Abdullah KL, Danaee M, Soh KL, Soh KG, Japar S. Stress and anxiety among mothers of premature infants in a Malaysian neonatal intensive care unit. J Reprod Infant Psychol 2019;37(2):193–205. doi: 10.1080/02646838.2018.1540861. Epub 2018 Nov 27. PMID:30480464.

18. van der Zee-van den Berg AI, Boere-Boonekamp MM, Groothuis-Oudshoorn CGM, Reijneveld SA. Postpartum depression and anxiety: a community-based study on risk factors before, during and after pregnancy. J Affect Disord. 2021;1:158–65. doi: 10.1016/j.jad.2021.02.062

19. Dennis CL, Falah-Hassani K, Brown HK, Vigod SN. Identifying women at risk for postpartum anxiety: a prospective population-based study. Acta Psychiatr Scand 2016; 134:485–93. doi: 10.1111/acps.12648

20. Lodhi FS, Rabbani U, Khan AA, Raza O, Holakouie-Naieni K, Yaseri M, et al. Factors associated with quality of life among joint and nuclear families: a population-based study. BMC Public Health 2021;21(1):234. doi: 10.1186/s12889-021-10265-2. PMID:33509153; PMCID: PMC7845136

21. Panchal DR. Mental health and psychological well-being among adolescents of joint and nuclear family. Int J Technol Res Eng 2013;7(4):431–4

22. Wenzel A, Haugen EN, Jackson LC, Brendle JR. Anxiety symptoms and disorders at eight weeks postpartum. J Anxiety Disord 2005; 19:295–311. PMID:15686858 DOI: 10.1016/j.janxdis.2004.04.001.

23. Hannon S, Gartland D, Higgins A, Brown SJ, Carroll M, Begley C, et al. Maternal mental health in the first year postpartum in a large Irish population cohort: the MAMMI study. Arch Womens Ment Health 2022;25(3):641–653. doi: 10.1007/s00737-022-01231-x. Epub 2022 Apr 29. PMID:35488067; PMCID: PMC9072451.

24. Reck C, Tietz A, Muller M, Seibold K, Tronick E. The impact of maternal anxiety disorder on mother-infant interaction in the postpartum period. PLOS ONE 2018; 13(5): e0194763.

25. Shensa A, Sidani JE, Dew MA, Escobar-Viera CG, Primack BA. Social media use and depression and anxiety symptoms: A cluster analysis. Am J Health Behav 2018;42(2):116–128. PMID 29458520.

